# Epidemiology of Inflammatory Bowel Disease in a Cohort of US Black Women

**DOI:** 10.1101/2022.07.14.22277547

**Authors:** Adjoa Anyane-Yeboa, Maame Araba E. Buadu, Hamed Khalili, Yvette Cozier

## Abstract

**Background and Aims:** Inflammatory bowel disease (IBD), including ulcerative colitis and Crohn’s disease are inflammatory diseases of the gastrointestinal tract. The incidence of IBD is increasing in minority populations; however, little is known about the epidemiology and disease characteristics of IBD in Black women.

**Methods:** Our study population included participants in the Black Women’s Health Study. Diagnosis of IBD was self-reported through the biennial questionnaires starting at baseline in 1995. We estimated the incidence of IBD according to age and geographic region. A follow up supplementary questionnaire was also sent to a subset of participants who reported diagnosis of IBD to evaluate the accuracy of self-reported diagnosis and to assess disease characteristics.

**Results:** Through December 31^st^ 2021, a total of 609 cases of IBD were reported, of which 142 were prevalent at baseline (prevalence = 0.24%) and 467 were incident (crude incidence rate = 33.2/100, 000 person-years). The incidence of IBD was highest in the <30 years age group and similar across geographic region. Among the participants who responded to the supplementary questionnaire, 62.1% had confirmed diagnosis of IBD.

**Conclusions:** In a large prospective cohort of US Black women, we found that the incidence of IBD was similar to previously published estimates in US White women. Future studies should focus on identifying risk factors for IBD in Black individuals in the US.

**What You Need to Know:** 

**Background:** The incidence of IBD is increasing in minorities.

**Findings:** The prevalence and incidence of IBD in the BWHS are higher than previously reported in other cohort studies and similar to those reported in US White women.

**Implications for Patient Care:** The burden of IBD in US Black women is high and similar to that of US White women.

## INTRODUCTION

Crohn’s disease (CD) and ulcerative colitis (UC) are chronic inflammatory diseases of the gastrointestinal tract with rising global incidence and prevalence^1,2^. In the United States (US), there are approximately 3 million individuals living with inflammatory bowel disease (IBD), which corresponds to a prevalence of approximately 1.3%^3^. A recent study from the Centers for Disease Control and Prevention (CDC) of Medicare beneficiaries over the age of 67 years from 2001 to 2018 reported the prevalence of CD and UC by race and ethnicity. As expected, the prevalence of CD and UC was highest among White individuals in the US, however the greatest increase in prevalence of IBD over this period was observed among non-Hispanic Black individuals. Similarly, the incidence of IBD has been reported to be highest in the non-Hispanic White population at 70.2/100,000 person-years (pyrs) compared to 24.9/100,000 person-years in the Black population^4^. The incidence, however, is increasing steadily in racial and ethnic minority groups. One recent study from Olmsted County, Minnesota showed an increase in the incidence of IBD by approximately 134% in the non-White population from 1970 to 2010, compared to an increase of only 39% in the White population^5^. In addition to the emerging data that suggest an increase in incidence and prevalence of IBD among Black individuals in the US, studies have also shown that IBD phenotype appears to be more aggressive among non-Hispanic Black individuals. Specifically, a number of studies have shown that a larger proportion of Black patients have penetrating, stricturing and perianal disease compared to White patients with CD^6-8^.

Furthermore, Black patients with UC and CD are more likely to undergo surgery compared to their White counterparts^6,7^. Despite the increasing burden of IBD in non-Hispanic Black individuals, no prior study has specifically examined the epidemiology of IBD among US Black individuals. Specifically, most prior epidemiologic studies of IBD to date have been in predominantly White populations such as the Nurses’ Health Study^9-12^, Olmstead County/Minnesota (USA)^5,13^, and Scandinavian cohorts^12^. To address this critical knowledge gap, we sought to build an infrastructure for studying the epidemiology of IBD in one of the few well characterized cohorts of Black individuals in the US, the Black Women’s Health Study (BWHS). Here, we describe our methods for ascertaining and validating IBD cases within BWHS and the preliminary results on the incidence of IBD in this cohort according to age and geographic region.

## MATERIALS AND METHODS

### Study Population

The BWHS is an ongoing prospective cohort study that was started in 1995 when a total of approximately 59,000 women, aged 21 to 69 years were enrolled through postal questionnaires sent to Black women’s professional organizations, *Essence* magazine subscribers, and friends and relatives of initial respondents^14,15^. Implied consent was determined by participants completing the questionnaires. On the baseline questionnaire (1995), participants were asked to provide information on demographics, medical and reproductive history, lifestyle factors (e.g., diet, physical activity), and use of certain medications such as oral contraceptives. Mailed biennial questionnaires update many of the data collected at baseline. Over 12 completed follow up cycles, cohort retention has averaged over 80%.

### Ascertainment and Validation of the Diagnosis of IBD

Starting with the baseline questionnaire in 1995, BWHS participants reported chronic conditions and age at diagnosis for a number of specific conditions (e.g., cancer, type-2 diabetes, myocardial infarction). Those not specified are reported under *“other serious illness”*. Cases of IBD were reported in this manner on all follow-up questionnaires, with the exception of 2013 and 2019. Women were specifically asked if they had ever been diagnosed with *“Crohn’s Disease (confirmed by biopsy)”* and the year of diagnosis on the 2013 follow-up questionnaire. On the 2019 questionnaire and every questionnaire thereafter, women were asked if they had ever been diagnosed with *“Crohn’s disease or ulcerative colitis”* and the year of first diagnosis.

To further confirm and characterize these cases, we designed a supplementary questionnaire for participants who reported a diagnosis of IBD. The supplementary questionnaire (**Appendix 1**) asked participants about symptoms related to their IBD, the course of their disease, number of hospitalizations, complications or surgeries, medications, family history and quality of life. Two gastroenterologists (HK and AAY) reviewed these supplementary questionnaires to confirm diagnosis of IBD. We mailed 107 supplementary questionnaires, and received 66 completed surveys for a 61.7% response rate. Among these, 49 participants confirmed the diagnosis of IBD and reported receiving ongoing treatment (74.2%). An additional 5 participants confirmed diagnosis of IBD but did not provide sufficient data in the supplementary questionnaire to assess the accuracy of the diagnosis. Finally, 12 participants reported being diagnosed with irritable bowel syndrome (n=10), ischemic colitis (n=1), and microscopic colitis (n=1).

### Statistical Analysis

Sample characteristics were summarized by calculating means and standard deviations. Chi-square tests of proportions compared characteristics according to IBD status (prevalent, incident). Person-time was estimated from baseline (1995) until the end of follow up on December 31^st^, 2021 (the end of the 2019 questionnaire cycle), date of diagnosis of IBD, date of last returned questionnaire or date of death, whichever came first. We estimated crude incident rates for IBD using the total number of reported cases diagnosed divided by total person-time. We compared crude incidence rates to those of other cohorts, based on their most recent publication^5,13,16,17^. We also calculated crude age- and geographic-specific incidence rates. Analysis was conducted using SAS v.9.4 Gary, NC). *P*-values less than 0.05 was considered statistically significant.

## RESULTS

### Baseline Characteristics of BWHS

At baseline in 1995, there were 58,973 women enrolled in the BWHS cohort. The median age of the cohort was 38 years (range=21-69 years), with the largest share in the 30-39 years category (33%). Most BWHS respondents had completed a Bachelor’s degree or had attended some college (24 and 36% respectively). There was broad geographic representation in the cohort with 27% from the Northeast, 31% from the South, 23% from the Midwest and 19% from the West. Approximately 56% worked in white collar occupations (**Table 1**).

**Table 1.** Baseline Characteristics of the Black Women’s Health Study

| **BWHS Cohort (N = 58,973)** |  |  |
| --- | --- | --- |
| **Characteristic** | **Number** | **%** |
| ***Age (years)*** |  |  |
| <30 | 12,809 | 22 |
| 30-39 | 19,577 | 33 |
| 40-49 | 16,440 | 28 |
| 50-59 | 7,235 | 12 |
| >60 | 2,912 | 5 |
| ***Education (years)*** |  |  |
| High school or less ($\leq 12$) | 11,396 | 19 |
| Some College (13 – 15) | 32,581 | 36 |
| Bachelor's (16) | 13,863 | 24 |
| Graduate ($>16$) | 12,413 | 21 |
| ***Geographic Region*** |  |  |
| Northeast | 16,107 | 27 |
| South | 18,030 | 31 |
| Midwest | 13,786 | 23 |
| West | 10,987 | 19 |
| ***Occupation*** |  |  |
| White collar | 33,071 | 56 |
| Non-White collar | 21,021 | 36 |
| Unemployed/Other | 3,232 | 5 |
| Never employed | 397 | 1 |
| Missing/Unknown | 1,251 | 2 |

### Comparison of incident and prevalent IBD cases in the BWHS

Among the 58,973 women in the BWHS cohort, 142 had IBD at the time of enrollment in the cohort for an overall prevalence of 0.24% at baseline (**Table 2**). Through the end of follow up in December of 2021, 467 individuals reported a diagnosis of IBD. As compared to prevalent cases, incident IBD cases were older and more likely to live in the Northeast. There were no differences in education, occupation, or whether they grew up in an urban or suburban neighborhood.

**Table 2.** Characteristics of reported incident and prevalent IBD cases in the Black Women’s Health Study

| Characteristic | Incident IBD<br>N = 467<br>n (%) | Prevalent IBD<br>N = 142<br>n (%) | P |
| --- | --- | --- | --- |
| **Age of diagnosis (years)** |  |  | 0.036 |
| <30 | 115 (25) | 17 (12) |  |
| 30-39 | 170 (36) | 59 (42) |  |
| 40-49 | 121 (26) | 44 (31) |  |
| 50-59 | 50 (11) | 18 (13) |  |
| >60 | 11 (2) | 4 (3) |  |
| **Education (years)** |  |  | 0.87 |
| High school or less ($\leq 12$) | 63 (14) | 19 (13) | |
| Some college (13 to 15) | 181 (39) | 51 (36) |  |
| Bachelor's (16) | 113 (24) | 40 (28) |  |
| Graduate (16+) | 109 (23) | 32 (23) |  |
| **Geographic Region** |  |  | 0.006 |
| Northeast | 133 (28) | 26 (18) |  |
| South | 129 (28) | 55 (39) |  |
| Midwest | 106 (23) | 38 (27) |  |
| West | 99 (21) | 22 (16) |  |
| **Occupation** |  |  | 0.213 |
| White collar | 278 (60) | 94 (66) |  |
| Blue collar | 158 (34) | 38 (27) |  |
| Unemployed/Never employed | 31 (6) | 10 (7) |  |
| **Where they lived up to age 18<sup>†</sup>** |  |  | 0.523 |
| Urban setting | 193 (46) | 66 (49) |  |
| Suburban setting | 72 (17) | 28 (21) |  |
| Rural or small town | 88 (21) | 25 (18) |  |
| Combination | 66 (16) | 16 (12) |  |

All variables taken from the baseline questionnaire in 1995
† Variable taken from the 1997 questionnaire

### Age and geographic distribution of reported incident IBD cases in the BWHS

Through the end of 2021, 58,973 women in BWHS contributed 1,405,348 person-years of follow up. The crude incidence rate for IBD was 33.2 cases per 100,000 person-years. **Figure 1a** shows the age-specific incidence of IBD in the BWHS as compared to other US cohorts. The highest incidence of IBD was in the <30 years age group at 49/100,000 person years. This finding was consistent with other US population cohorts that have shown the highest incidence of IBD in adults less than 30 years old (**Figure 1a)**. Our estimates, however, were higher in the 30-59 years age group in comparison to other cohorts. We performed a sensitivity analysis where we applied the 62.1% confirmation rate to our incidence estimates. This adjustment brought the crude incidence of IBD in the BWHS into alignment with other US cohorts across all age groups. The crude incidence rates were similar across different US geographic regions and varied from 30 cases/100,000 person-years in the southern region to 38 cases/100,000 person-years in the western region (**Figure 1b**).

**Figure 1:**
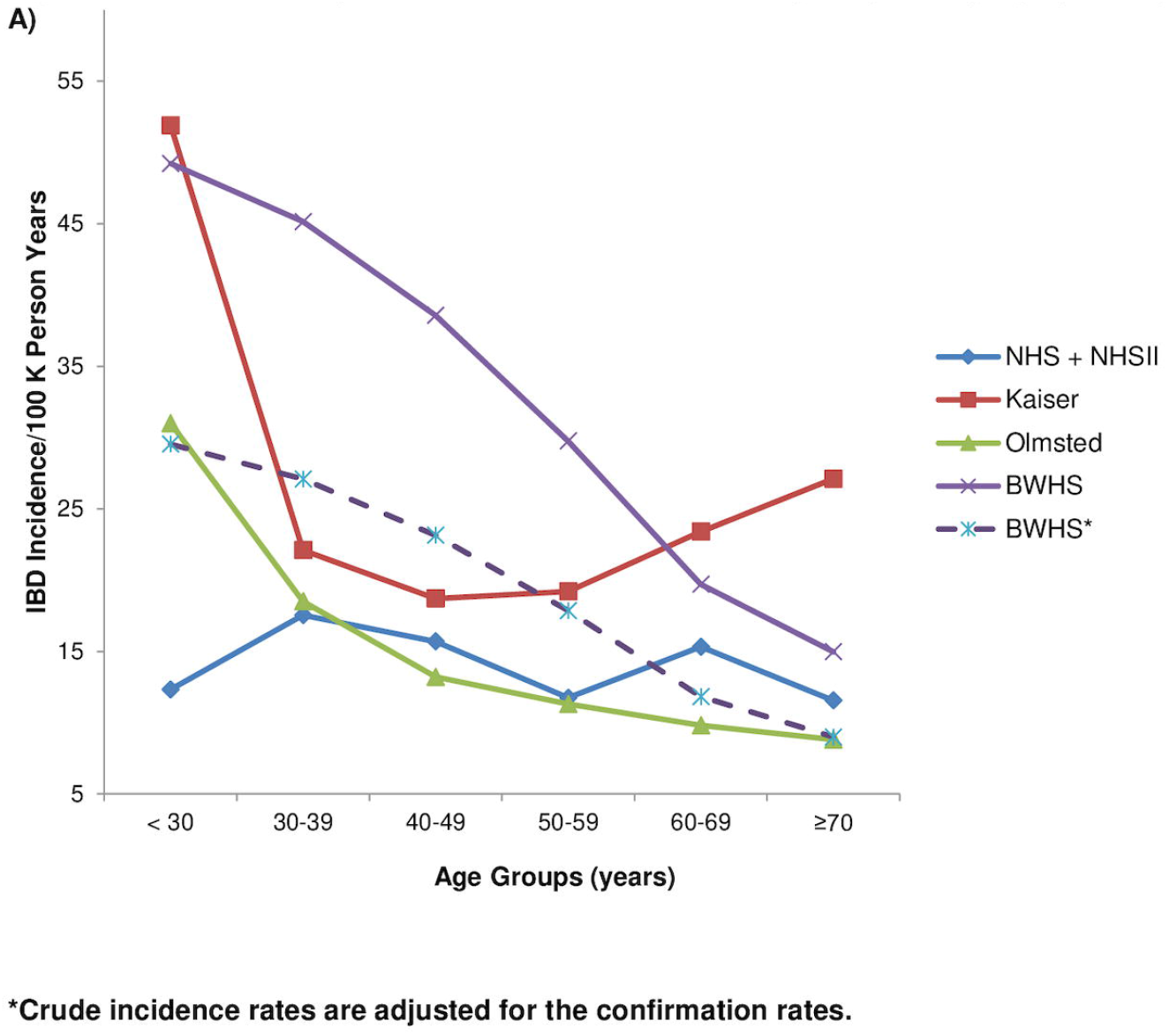
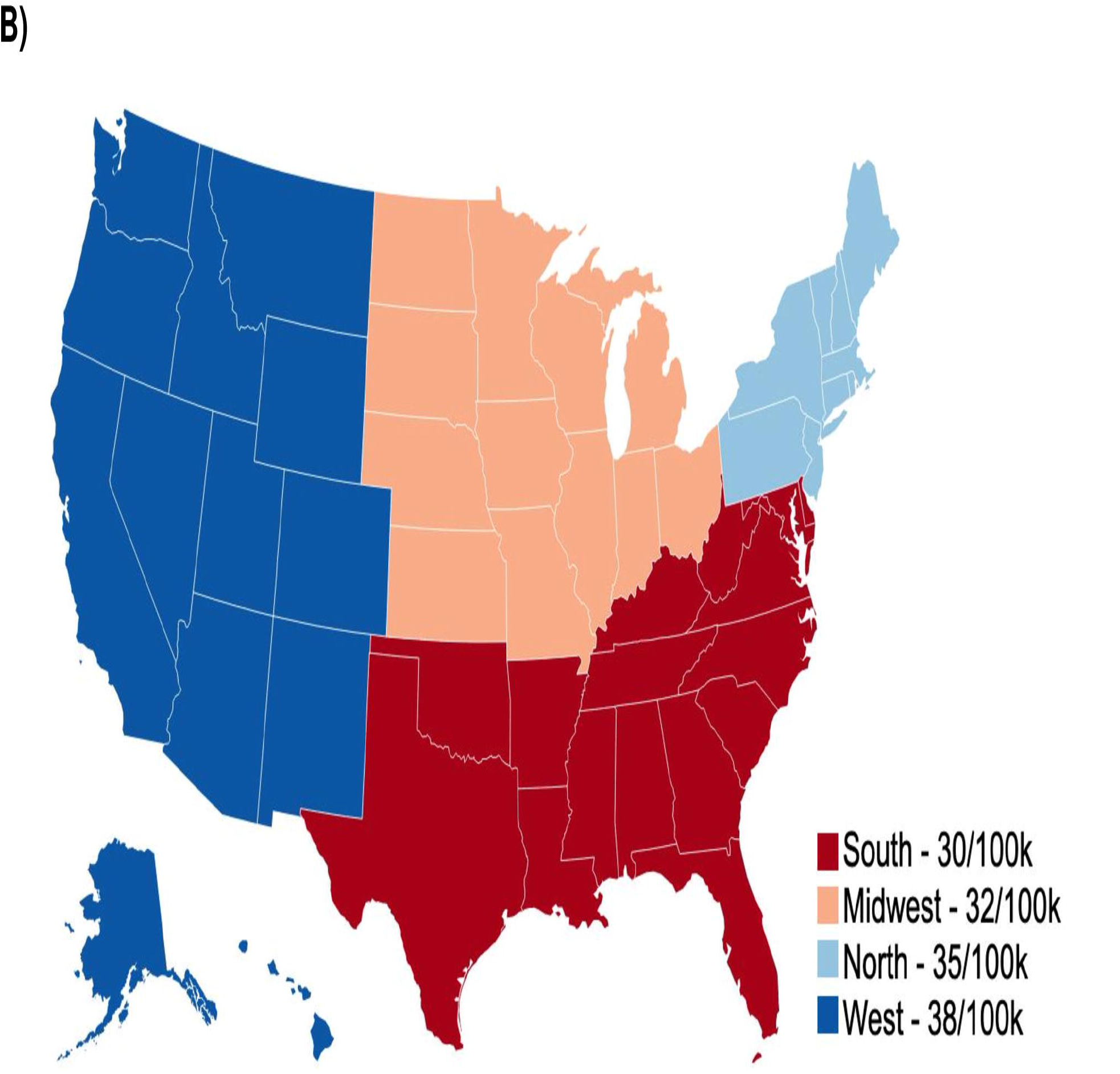
Incidence of reported IBD in the BWHS according to age and geographic region.

### Characteristics of IBD cases who returned the supplementary questionnaire in the BWHS

Among participants who returned the supplementary questionnaire, CD was the most common subtype at 45%, followed by UC at 37%, and 9% IBD unclassified (**Table 3**). The mean age at diagnosis was 36.7 years (range: 14-61 years) for UC cases, and 31.9 years (range: 18-56 years) for CD cases. Of the confirmed cases, 26% had previously been hospitalized for IBD, with 72% of those hospitalized having had 2 or more hospitalizations. Approximately 33% (n=22) reported history of an IBD-related surgery, 43% reported a history of extraintestinal manifestations, and 44% reported history of complications of IBD including stricture, fistula or perianal disease.

**Table 3.** Characteristics of Confirmed Cases of IBD in the Black Women’s Health Study

| Characteristic |  |  |
| --- | --- | --- |
| IBD subtype | Number (%) | Age at diagnosis, years mean (SD; range) |
| UC | 18 (37) | 36.7 (14.5; 14 - 61) |
| CD | 22 (45) | 31.9 (9.7; 18 – 56) |
| IBD-U | 9 (18) | 44.1 (11.6; 25 – 60) |
|  | Number (%) |  |
| Hospitalizations for IBD (yes) | 18 (26) |  |
| 1 | 5 (28) |  |
| 2 or more | 13 (72) |  |
| Complications for IBD* (yes) | 31 (44) |  |
| 1 | 16 (52) |  |
| 2 or more | 15 (48) |  |
| IBD-related surgery (yes) | 23 (33) |  |
| 1 | 17 (74) |  |
| 2 or more | 6 (26) |  |
| Medications Used |  |  |
| Corticosteroids | 29 (41) |  |
| 5-ASA | 32 (46) |  |
| Biologics | 12 (17) |  |
| Immunosuppressants | 10 (14) |  |
| EIM (co-morbid conditions) (yes) | 30 (43) |  |
| 1 | 14 (47) |  |
| 2 or more | 16 (53) |  |
| Family history of IBD (yes) | 11 (16) |  |
| Self-rated Health |  |  |
| Excellent | 11 (16) |  |
| Good | 30 (43) |  |
| Fair | 15 (21) |  |
| Poor | 3 (4) |  |
| Very poor | 1 (1) |  |
| Not reported | 10 (14) |  |
| *Complications for IBD includes the following categories: 1) stricture/blockage/bowel obstruction; 2) fistula; and 3) perianal disease. |  |  |

In terms of medication history, 41% reported ever being on corticosteroids and 46% on 5-aminosalicylate medications. Only 17% had ever been on biologics and 14% on immunosuppressants (**Table 3**). Majority of respondents rated their overall health as fair (21%), good (43%), and excellent (16%).

## DISCUSSION

Here we present the preliminary results on the epidemiology of IBD in a well-established cohort of Black women, BWHS. We found that the crude incidence rate of reported IBD and the age distribution were similar to those of other population-based cohorts that have primarily included White participants. The incidence rates of reported IBD were similar across different geographic region in the US. We further demonstrated that the confirmation rate of IBD in a subset of participants was around 60% and similar to those of other population-based cohorts. Lastly, a significant proportion of IBD cases who provided additional information on their disease reported having experienced complications or required surgery.

The incidence of IBD is increasing in the US Blacks. Historically IBD has been thought of as a disease that impacts primarily White individuals of European descent, but in recent decades the burden of IBD has increased significantly in racial and ethnic minorities^5^. In a national cohort of 25.1 million Medicare fee-for-service beneficiaries, the age-adjusted prevalence of IBD was 1.2% overall, 1.4% in non-Hispanic White individuals, and 0.5% in Black individuals^18^. Similarly, in a population-based cohort study from Olmsted, Minnesota, the incidences of IBD was noted to be 21.6 cases per 100,000 person-years in White individuals and 13 per 100,000 person years in minorities^5^. The authors noted that the incidence of IBD in minority individuals had increased by 134% from 1970 to 2010 compared to only 30% in White individuals^5^. Interestingly in our study, we found a higher incidence of 33.2/100,000 person-years in US Black women, compared to what has been shown in other US-based cohorts.

Racial and ethnic differences in IBD phenotype, complications, and medication usage have been reported in prior studies. A study from a tertiary care center noted racial differences in IBD phenotype between Black and White patients with IBD. The authors noted that Black patients with IBD were more likely to experience extraintestinal manifestations in the form of arthralgia as compared to White patients with IBD^7^. In addition, Black patients with UC in this cohort had a higher prevalence of ankylosing spondylitis and sacroiliitis than White patients^7^. In a recent systematic review, the authors concluded that Black patients with CD are more likely to have complicated CD defined as having perianal or penetrating disease as compared to White patients with CD^6^. These findings are in line with our observation that nearly half of patients with IBD in BWHS experienced a disease complication.

The cause of the rising incidence of IBD in US Black individuals in unclear. One explanation may be underdiagnosis. One recent study noted that Black patients who presented with iron deficiency anemia and chronic diarrhea, which may represent a diagnosis of IBD, were less likely to receive an appropriate workup^19^. In recent decades there has been an increased awareness around IBD in minority groups which may have contributed to then more recent rise in recognition of disease in this population. Another potential source of disparity is rooted in the social determinants of health and structural racism which can impact where individuals live, shape their access to healthy food options, ability to exercise and exposure to adverse behaviors like tobacco usage^20^. For example, some epidemiologic studies of IBD have noted that diet has an impact on the incidence of IBD, and specifically diets high in trans-unsaturated fat, red meat and low in fiber may increase the risk of developing IBD^10,11,21^. Data also suggest that dietary consumption patterns differ according to race and ethnicity and may therefore contribute to rise in IBD incidence among US Black individuals^22,23^.

There are a few limitations of our study that are worth noting. First, our study is a cohort of exclusively US Black women and it is unclear if the findings would be generalizable to Black men. There could be potential sex differences in the incidence of IBD and thus additional studies including Black men are needed. Secondly, we note that our crude incidence rates are based on self-reported diagnosis of IBD and may therefore overestimate the true rates. However, in our validation study of a subgroup of participants reporting IBD, we noted that the confirmation rate exceeded 60%. When we applied the confirmation rate to our incidence estimates, our results were similar to those from other population cohorts that largely included White populations.

Additionally, IBD may have also been underreported in our cohort. Lastly, our study population consisted of predominantly educated Black women with approximately 80% having education beyond high school. This study may therefore underrepresent the 15% who have not completed high school; however our population is in line with what is seen broadly in Black individuals in the US, with rates of high school completion in this population at 84.5% in 2011 and 90.3% in 2021^24^.

Our study has several strengths. We present data on the incidence and characteristics of IBD from a large nationally representative cohort of US Black women. Prior epidemiologic studies in IBD consist of predominantly White individuals, thus our findings present unique insight into the epidemiology of IBD in a cohort of Black individuals. In addition, we conducted a pilot study to further demonstrate the validity of self-reported diagnosis of IBD in our cohort.

We present a descriptive study of the incidence, prevalence and characteristics of IBD in a cohort of Black women in the US. Our preliminary analyses demonstrate that the incidence of IBD in Black women rivals the estimates from cohorts that consist of predominantly White participants. Additionally, similar to previously published data, we note that incidence of IBD in Black women is highest in young adulthood. Lastly, our data demonstrate the significant burden of IBD among US blacks as demonstrated by high rate of complications. Future studies are needed to further investigate the role of individual dietary and lifestyle factors and social and contextual factors on the rising incidence of IBD in the Black population in the US.

## Data Availability

Data underlying the study cannot be made publicly available due to ethical concerns about patient confidentiality. Data will be made available to qualified researchers on request to.

## Author Contributions

AAY data curation and validation, investigation, writing-original draft, writing – review and editing, visualization; MAEB writing-original draft, writing- review and editing; YC conceptualization, methodology, validation, formal analysis, investigation, data curation, writing- review & editing, supervision. HK conceptualization, methodology, validation, investigation, writing- review & editing, supervision

