## Supplemental material for "Epidemiology of Inflammatory Bowel Disease in a Cohort of US Black Women"

### Crohn's IBD Questionnaire

**1. Please write in your age and date of birth.**

| Age | Month (example: June = 06) | Day | Year |
|---|---|---|---|
| | | | 1 9 |

**2. Which of the following have you <u>ever</u> been diagnosed with (check all that apply):**

| | Age at diagnosis |
|---|---|
| ○ Ulcerative Colitis | |
| ○ Crohn's disease | |
| ○ Inflammatory Bowel Disease | |
| ○ Microscopic Colitis | |
| ○ Irritable Bowel Syndrome | |

**3. <u>Including your first episode</u>, have you experienced any of the following symptoms related to your ulcerative colitis, Crohn's disease, inflammatory bowel disease, or Microscopic Colitis (Collagenous or Lymphocytic)? Please indicate the longest duration of each symptom. (Check all that apply).**

| Symptom | Duration of symptoms: Less than 2 weeks | 2 to 3 weeks | 4 weeks or greater |
|---|---|---|---|
| Abdominal pain | ○ | ○ | ○ |
| Diarrhea | ○ | ○ | ○ |
| Rectal bleeding/ blood in stool (not due to hemorrhoids or constipation) | ○ | ○ | ○ |
| Fever greater than 100.5°F | ○ | ○ | ○ |
| Fatigue or lack of energy | ○ | ○ | ○ |
| Difficulty sleeping | ○ | ○ | ○ |
| Nausea | ○ | ○ | ○ |
| Vomiting | ○ | ○ | ○ |
| Mouth sores | ○ | ○ | ○ |
| Back pain | ○ | ○ | ○ |
| Night sweats | ○ | ○ | ○ |
| Decreased appetite | ○ | ○ | ○ |
| Weight loss | ○ | ○ | ○ |

**4. Which of the following profiles most accurately reflects the course of your disease?**

Initial flare, then inactive ○

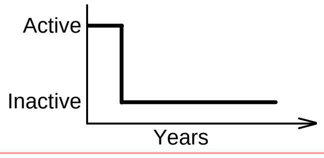


Chronically active ○

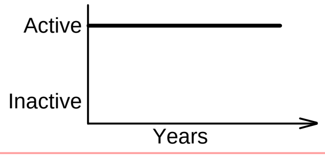


Less than 1 flare/ year ○

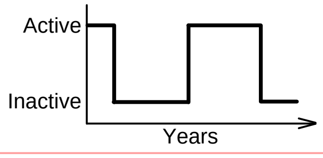


One or more flares/year ○

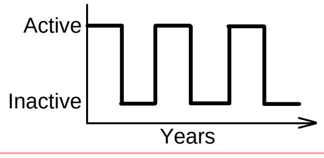


**5. IN THE PAST SIX MONTHS, my disease has been:**

- ○ Constantly active, giving me symptoms every day
- ○ Often active, giving me symptoms most days
- ○ Sometimes active, giving me symptoms some days (for instance, 1 - 2 days per week)
- ○ Occasionally active, giving me symptoms 1 - 2 days per month
- ○ Rarely active, giving me symptoms on a few days the past 6 months
- ○ I was well in the past 6 months, what I consider a remission or absence of symptoms

*Next page, please.* →

6. **Since your initial diagnosis, how many times have you been hospitalized for colitis or Crohn's disease?** ☐☐

| | Name of hospital | month | year |
|---|---|---|---|
| 1) | | / | |
| 2) | | / | |
| 3) | | / | |

***For those with a Microscopic Colitis Diagnosis Only, please skip to question 10.*** →

7. **Have you ever had any of the following complications or surgeries? (Check all that apply).**

| | Date first diagnosed month / year | | Date first diagnosed month / year |
|---|---|---|---|
| ○ Perianal or perirectal fistula | / | ○ Resection or removal of small intestine | / |
| ○ Fistula between intestine and vagina or bladder | / | ○ Partial colectomy | / |
| ○ Fistula between intestine and skin (draining) | / | ○ Total colectomy | / |
| ○ Fistula between intestines | / | ○ Ileostomy | / |
| ○ Stricture (narrowing) of the intestine or colon | / | ○ Ileal or J-pouch | / |
| ○ Bowel obstruction | / | ○ Appendectomy | / |
| ○ Abscess (collection of pus) in the abdomen | / | ○ Gallbladder surgery | / |

8. **How many surgeries (NOT including endoscopies) have you had for ulcerative colitis or Crohn's disease?** ☐☐

| | Type of surgery | Name of hospital | month | year |
|---|---|---|---|---|
| 1) | | | / | |
| 2) | | | / | |
| 3) | | | / | |

9. **Has a physician ever diagnosed any of the following conditions in you? (Check all that apply).**

| | Date first diagnosed month / year |
|---|---|
| ○ Uveitis (eye inflammation) | / |
| ○ Iritis (eye inflammation) | / |
| ○ Pyoderma gangrenosum (ulcerating skin disease) | / |
| ○ Erythema nodosum (painful red bumps) | / |
| ○ Colon cancer | / |
| ○ Precancerous lesions in the colon (e.g. polyps or dysplasia) | / |
| ○ Pouchitis (inflammation in ileal or J-pouch) | / |
| ○ Arthritis or joint swelling due to colitis/Crohn's | / |
| ○ Ankylosing spondylitis | / |
| ○ Primary sclerosing cholangitis | / |
| ○ Blood clot, venous thrombosis or embolism | / |
| ○ Kidney stones | / |

***Next page, please.*** →

**10. For your ulcerative colitis, Crohn's disease, inflammatory bowel disease or Microscopic Colitis (Collagenous or Lymphocytic), what treatments have you received or have you been receiving and how did the following medications make you feel? (Check all that apply).**

***Instructions:***

***Full recovery:*** *medication made me feel completely better*
***Partial recovery:*** *medication made me feel a little (partially) better*
***No recovery:*** *medication did not make me feel any better*
***Felt worse:*** *had to stop taking because of side-effects*

| Medication | Taken: At first episode | Taken: Ever | Date started: month | Date started: year | Date stopped: month | Date stopped: year | What was your response to the medication? Full recovery | Partial recovery | No recovery | Felt worse |
|---|---|---|---|---|---|---|---|---|---|---|
| Oral sulfasalazine | ○ | ○ | | | | | ○ | ○ | ○ | ○ |
| Oral mesalamine (e.g., Dipentum, Asacol, Pentasa, Colazal, Lialda, Apriso, Delzicol) | ○ | ○ | | | | | ○ | ○ | ○ | ○ |
| Intravenous steroids (e.g., solumedrol, hydrocortisone) | ○ | ○ | | | | | ○ | ○ | ○ | ○ |
| Oral prednisone | ○ | ○ | | | | | ○ | ○ | ○ | ○ |
| Oral budesonide (e.g., Entocort) or Budenoside MMX (e.g. Uceris) | ○ | ○ | | | | | ○ | ○ | ○ | ○ |
| Mesalamine suppository or enema (e.g., Canasa, Rowasa) | ○ | ○ | | | | | ○ | ○ | ○ | ○ |
| Steroid suppository or enema | ○ | ○ | | | | | ○ | ○ | ○ | ○ |
| Antibiotics | ○ | ○ | | | | | ○ | ○ | ○ | ○ |
| Cyclosporine | ○ | ○ | | | | | ○ | ○ | ○ | ○ |
| 6-mercaptopurine (6MP) or azathioprine (Imuran) | ○ | ○ | | | | | ○ | ○ | ○ | ○ |
| Methotrexate | ○ | ○ | | | | | ○ | ○ | ○ | ○ |
| Infliximab (Remicade) | ○ | ○ | | | | | ○ | ○ | ○ | ○ |
| Certolizumab (Cimzia) | ○ | ○ | | | | | ○ | ○ | ○ | ○ |
| Adalimumab (Humira) | ○ | ○ | | | | | ○ | ○ | ○ | ○ |
| Natalizumab (Tysabri) | ○ | ○ | | | | | ○ | ○ | ○ | ○ |
| Vedolizumab (MLN0002) | ○ | ○ | | | | | ○ | ○ | ○ | ○ |
| Golimumab (Simponi) | ○ | ○ | | | | | ○ | ○ | ○ | ○ |
| Other | ○ | ○ | | | | | ○ | ○ | ○ | ○ |

Please specify

**You may add additional comments here about your use of medications or treatments:**

***For those with a Microscopic Colitis Diagnosis Only, please skip to question 14.*** →

***Next page, please.*** →

**11. Not counting half siblings, how many siblings do you have?** Brothers [ ][ ] Sisters [ ][ ]

**12. How many biological children do you have?** Sons [ ][ ] Daughters [ ][ ]

**13. Do you have a family history of Crohn's (CD), ulcerative colitis/proctitis (UC) or inflammatory bowel disease (IBD) not otherwise specified (NOS)?**

| | Type of disease | | | Age of first diagnosis | | | | | | | | |
|---|---|---|---|---|---|---|---|---|---|---|---|---|
| Relative | CD | US | IBD (NOS) | <20 | 20-29 | 30-39 | 40-49 | 50-59 | 60-69 | 70-79 | >80 | Don't know |
| Mother | ○ | ○ | ○ | ○ | ○ | ○ | ○ | ○ | ○ | ○ | ○ | ○ |
| Father | ○ | ○ | ○ | ○ | ○ | ○ | ○ | ○ | ○ | ○ | ○ | ○ |
| *Do not include half siblings* | | | | | | | | | | | | |
| Brother 1 | ○ | ○ | ○ | ○ | ○ | ○ | ○ | ○ | ○ | ○ | ○ | ○ |
| Brother 2 | ○ | ○ | ○ | ○ | ○ | ○ | ○ | ○ | ○ | ○ | ○ | ○ |
| Sister 1 | ○ | ○ | ○ | ○ | ○ | ○ | ○ | ○ | ○ | ○ | ○ | ○ |
| Sister 2 | ○ | ○ | ○ | ○ | ○ | ○ | ○ | ○ | ○ | ○ | ○ | ○ |
| *Biological children only* | | | | | | | | | | | | |
| Son 1 | ○ | ○ | ○ | ○ | ○ | ○ | ○ | ○ | ○ | ○ | ○ | ○ |
| Son 2 | ○ | ○ | ○ | ○ | ○ | ○ | ○ | ○ | ○ | ○ | ○ | ○ |
| Daughter 1 | ○ | ○ | ○ | ○ | ○ | ○ | ○ | ○ | ○ | ○ | ○ | ○ |
| Daughter 2 | ○ | ○ | ○ | ○ | ○ | ○ | ○ | ○ | ○ | ○ | ○ | ○ |
| *Additional relatives* | | | | | | | | | | | | |
| [ ] | ○ | ○ | ○ | ○ | ○ | ○ | ○ | ○ | ○ | ○ | ○ | ○ |
| [ ] | ○ | ○ | ○ | ○ | ○ | ○ | ○ | ○ | ○ | ○ | ○ | ○ |

**14. How would you rate your health today with regard to your ulcerative colitis, Crohn's disease, inflammatory bowel disease, or Microscopic Colitis (Collagenous or Lymphocytic)?**

Excellent ○ Good ○ Fair ○ Poor ○ Very poor ○

**You may add additional comments here :**

[ ]
